# Evaluation of a clinical decision support system for detection of patients at risk after kidney transplantation

**DOI:** 10.1101/2022.05.12.22275019

**Authors:** Roland Roller, Manuel Mayrdorfer, Wiebke Duettmann, Marcel G. Naik, Danilo Schmidt, Fabian Halleck, Patrik Hummel, Aljoscha Burchardt, Sebastian Möller, Peter Dabrock, Bilgin Osmanodja, Klemens Budde

## Abstract

Patient care after kidney transplantation requires integration of complex information to make informed decisions on risk constellations. Many machine learning models have been developed for detecting patient outcomes in the past years. However, performance metrics alone do not determine practical utility. Often, the actual performance of medical professionals on the given task is not known. We present a newly developed clinical decision support system (CDSS) for detection of patients at risk for rejection and death-censored graft failure. The CDSS is based on clinical routine data including 1516 kidney transplant recipients and more than 100 000 data points. Additionally, we conduct a reader study to compare the performance of the system to estimations of physicians at a nephrology department with and without the CDSS. Internal validation shows AUC-ROC scores of 0.83 for rejection, and 0.95 for graft failure. The reader study shows that although the predictions by physicians converge towards the suggestions made by the CDSS, performance in terms of AUC-ROC does not improve (0.6413 vs. 0.6314 for rejection; 0.8072 vs. 0.7778 for graft failure). Finally, the study shows that the CDSS detects partially different patients at risk compared to physicians without CDSS. This indicates that the combination of both, medical professionals and a CDSS might help detect more patients at risk for graft failure. However, the question of how to integrate such a system efficiently into clinical practice remains open.

## Introduction

Kidney transplantation is the treatment of choice for patients with end-stage kidney disease (ESKD) although it requires lifelong post-transplant care^1,2^. Graft failure is often multifactorial^3,4^, therefore it is important to continuously account for a diverse set of potentially detrimental events in clinical care, depending on individual patient risk profiles. The heterogeneity of causes leading to graft failure makes it very challenging to predict the course of a transplant life and finally graft failure. Given the high number of graft failures affected by over-immunosuppression (infections, drug toxicity, cancer) and under-immunosuppression (rejection), adjustment of immunosuppressive treatment is one of the most powerful tools in clinical practice^3-5^. However, it is often not clear how to interpret the current risk profile due to an overwhelming amount of data to be integrated for decision-making. This dilemma is further enhanced by the lack of time in clinical routine. Therefore, clinical decision support systems able to integrate and interpret the often highly complex status of a kidney transplant recipient are an interesting option to mitigate this problem. In recent years, an increasing number of machine learning (ML) solutions have been developed to support medical professionals. However, many publications revolve around novel ML models with the goal of outperforming baselines and pushing the boundaries in terms of better performance^6,7^. Only a few approaches go beyond the pure improvement of ML models and provide detailed technical analyses or insights about how and what the model has learnt^8,9^. Unfortunately, in most cases the system is not evaluated together with the end user. This renders interpretation of the actual impact in clinical routine difficult. In order to improve medical care in the real world, ML models not only have to be accurate and precise, they also need to be embedded in the patient journey and accepted by medical professionals. In this work, we present a Clinical Decision Support System aiming to detect in advance patients at risk of a) rejection, and b) death-censored graft failure, to occur within the next 90 days. Additionally, we evaluate the system together with medical professionals.

## Methods

For simplicity, we refer to death-censored graft failure as graft failure, physician as MD (medical doctor), ML-based clinical decision support system as AI, and physicians with AI support as MD+AI.

### Data

Baseline for this work is TBase^10^, a database designed for kidney transplant recipients (KTR), implemented over 20 years ago at Charité. As patients are supposed to receive a follow-up at the transplant center 3-4 times a year, TBase includes fine-grained information about the patients over many years including demographics, laboratory data, medication, medical notes, diagnoses, radiology, and pathology reports. Death-Censored Graft Failure is defined as the initiation of renal replacement therapy (dialysis or re-transplantation). Graft loss due to death with a functioning graft is not included in the current work. Rejection is defined according to the Banff 2017 classification^11^, as previously described^3^.

### Data Selection, Enrichment and Cohort Generation

The complete risk prediction scenario is built up around data points (DP), which describe the particular moment when new data about a patient is inserted in the database. If one of the endpoints occurs within the target prediction window of 90 days, the DP will be labeled as true, otherwise as false. According to those DPs, patient information (features) which are available at this moment, are extracted (e.g., the new and the previous lab values, or the current diagnoses) and used as input information of our model. Moreover, we enrich the data by additional information, such as mean scores or gradients of successive values. Each model uses about 300 different features, consisting of structured information (e.g., vital parameters, lab values, medication) and bag-of-word features (single word features taken from findings or diagnoses). Table 1 provides an overview of the most relevant features for each model. A more detailed overview of the features used, is provided in table S6.

**Table 1:** Overview of the most relevant features (top 5) of each model (global features), including its importance (Imp.)

| *Rejection* |  | *Graft Loss* |  |
| --- | --- | --- | --- |
| *Feature* | *Imp.* | *Feature* | *Imp.* |
| *Serum creatinine (lab value)* | *12.78* | *last transplantation (months)* | *35.12* |
| *last transplantation (months)* | *7.66* | *# of transplantations* | *23.76* |
| *rejections in last 180 days* | *7.26* | *Serum creatinine (lab value)* | *8.99* |
| *last transplantation (days)* | *6.69* | *last transplantation (days)* | *3.04* |
| *# of lab values last 60 days* | *3.18* | *eGFR (lab value)* | *2.26* |

Next, data is filtered to generate a meaningful, reliable, valid and realistic dataset: Only data points with a follow-up data point within the next 15 to 180 days are used. This filter has been implemented to exclude gaps in patient follow-up ensuring reliability of endpoint evaluation. The resulting dataset is referred to as “cohort”. The cohort includes 1516 different patients, with a mean of 67.89 data points (moments during transplant life with new data input) per patient. The cohort is then cleaned and divided into a training, development, and test dataset for ML. An overview about additional data characteristics as well as the exclusion criteria is provided in the supplementary material.

### Clinical Decision Support System

The ML component relies on Gradient Boosted Regression Trees (GBRTs). In comparison to neural methods, GBRTs can be quickly trained, and therefore quickly modified and further optimized, without the need of a strong computer cluster. Also, tree-based methods are well established in the context of CDSS, and can be also easier understood by non-experts, thus exhibiting some sort of transparency.

The resulting dataset is strongly unbalanced, as it contains a larger number of negative instances compared to positive ones. That means, in most cases an endpoint does not occur in the target period for the given data point (Table S3). As a large portion of negative data can influence the quality of the classifier, and slows down the training, negative samples are randomly down-sampled within the training split. Controlled upsampling (SMOTE)^12^, and controlled downsampling (NCA)^13^ did not lead to any significant improvements. For the final setup, a training ratio of 1:3 was chosen, as it showed the most promising results during the initial experiments.

For the internal validation, we randomly assigned 70% of the patients into the training, 15% into the development and 15% into the test split. This step is repeated 50 times for the cross-validation. That way, patients within training, development and test data always change. Therefore, reported mean scores and 95% CI (confidence interval) provide a good approximation of our model.

For the reader study instead, data is prepared differently: First, the test set is defined and all patients of the test set removed from the cohort. The test set contains 120 patients, which was the sample size calculated in the power analysis shown in supplementary item K. As endpoints have a low frequency, and to ensure that a sufficient number of events occur, data points are selected randomly, but in a controlled way. Each endpoint occurs at least 20 times in the reader study test set. Then, the remaining patients with their data points are split into a training and development set, using a split of 80% and 20%. Note, as opposed to the AI system, the physician can access the complete patient history, including text notes, medical reports, and a longer time range than the last two entries (e.g., laboratory tests) of the last year. The physicians in the reader study are informed that the endpoint frequency might not reflect real-world conditions but ^14^are not informed about the exact distribution.

The final ML model relies on GBRTs implemented in python using scikit-learn, with 300 estimators, a learning rate of 0.1, max-depth of 3, and a random-state of 0. The model is trained on a Ubuntu 18.04.3 LTS server with a Intel Core i9-7900X 3.30GHz. The training of the model with ca. 10k training examples take about 2.5 minutes. This would describe one cross-validation step in our first experiment. The model with technical descriptions can be made available on request.

### Dashboard

To provide an informative decision support system, a dashboard was developed including the following information: a) current risk score, b) development of the risk score over time within a graph, c) categorization of the risk scores into a traffic light system (green, yellow, red), as well as d) presentation of relevant features which influence the decision of the risk score. For each given data point, two dashboard-graphs (including the additional information) are generated, one for each endpoint.

Figure 1 presents an example of the dashboard, presenting the time on the x-axis and the risk score on the y-axis. All data points of the last year, including corresponding risk scores, are visualized in the graph. The graph itself is divided into different zones, a green zone indicating a low risk, a yellow zone which indicates a higher risk, and a red zone which indicates the highest risk. Each zone is defined by a threshold, which was generated on the development set by identifying the optimal F-Scores - F2 score for the border to the yellow zone, and F0.5 to indicate the red zone.

**Figure 1:**
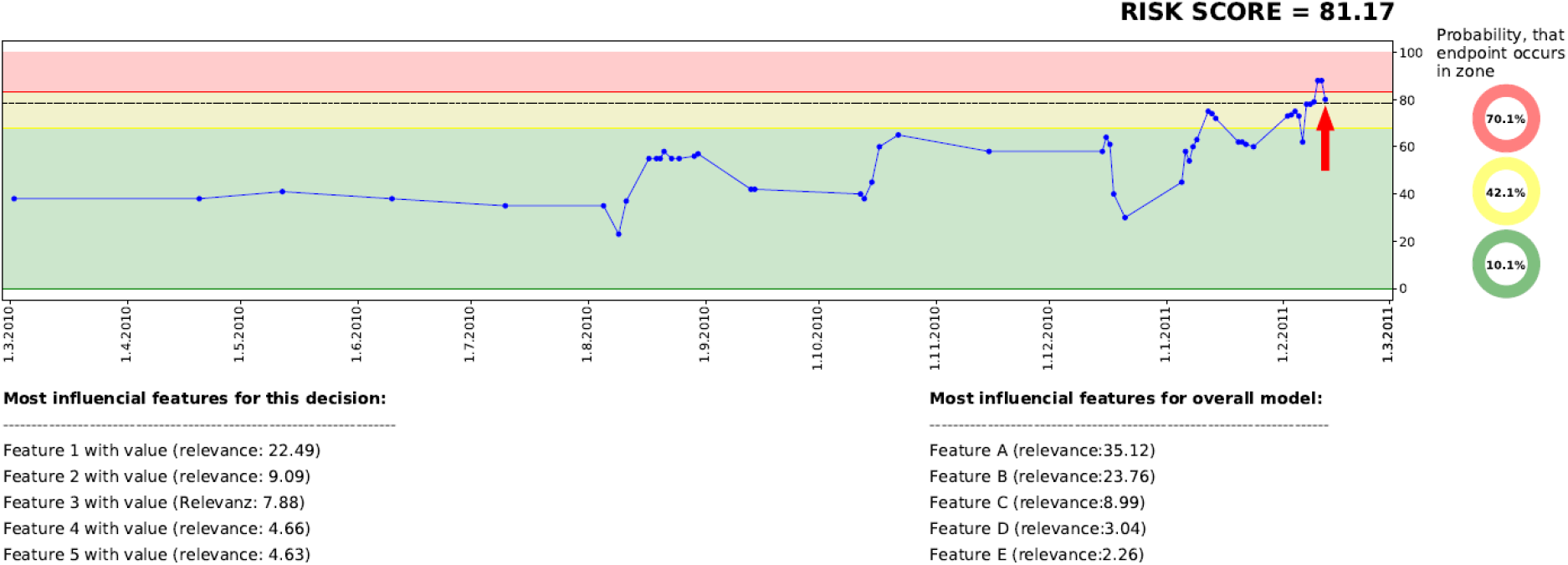
Visualization of the dashboard, including historic risk scores, a traffic light system, a well as a model and a decision-based explanation.

### Reader Study Design

Overall, eight physicians, four junior MDs who have not completed their specialization and four senior MDs with specialization in internal medicine or nephrology, participate in the reader study (for participant demographics see Table S5 in the supplementary material). MDs were to examine all information available about the kidney transplant recipient in the database and to forecast the two endpoints, once without and once with the help of AI. Each MD examines 15 different DPs with the corresponding patient case history without CDSS, and then 15 other DPs of different patients with CDSS. The DPs are randomly assigned to the physicians in both parts of the study, so that no physician assesses the same patient twice. Before starting the second part of the experiment, each physician receives a small tutorial (see K1 supplementary material) to understand the dashboard of the CDSS. In the first step, the MD receives a data point of the cohort and estimates how likely, in terms of probability (0% to 100%), each endpoint might occur in our target period (90 days). In the second part, the dashboard with the AI prediction is presented simultaneously with patient data (MD+AI). In both rounds, each MD has up to 30 minutes time to study the de-identified medical history of the patients. Physicians were provided with the exact endpoint definitions.

### Significance Tests

In order to examine the significance of the different ROC (Receiver Operating Characteristic) scores in our study, the implementation of DeLong^14^ is applied (paired, one-tailed, in case of Table 3; and unpaired, one-tailed in case of Table 4). To explore the significance of the influence of the AI system, we use a two-tailed t-test implemented in SciPy.

## Results

### Internal Validation

The results of the internal validation are presented in Table 2. Considering the prediction period of 90 days, our model shows an AUC-ROC (Area Under the Curve of the Receiver Operating Characteristic) score of 0.83 and 0.95 for rejection and graft failure, respectively. The results also show that model performance shows only slight changes when extending the prediction period to 180 or 360 days.

**Table 2:** Risk prediction results in terms of AUC-ROC including 95% CI during internal validation, using a resampling approach with 50-fold cross validation on retrospective data.

| *Endpoint* | *days* | *AUC-ROC* |
| --- | --- | --- |
| ***Rejection*** | 90 | 0.832 (0.04), 95% CI [0.771, 0.903] |
|  | 180 | **0.824** (0.03), 95% CI [0.769, 0.892] |
|  | 360 | **0.811** (0.04), 95% CI [0.736, 0.868] |
| ***Graft Failure*** | 90 | 0.945 (0.02), 95% CI [0.901, 0.970] |
|  | 180 | **0.947** (0.01), 95% CI [0.924, 0.969] |
|  | 360 | **0.953** (0.01), 95% CI [0.934, 0.972] |

### Reader Study

For the reader study, a new ML model is trained from scratch, using the configuration of the internal validation, but using a new split of training, development, and test data, as described above. The predictions of the ML model on the test set are used as CDSS for the physicians (MD+AI).

### AI vs MD vs AI+MD

The results of the reader study are presented in Table 3 and Figure 2. The results show that AI outperforms MDs (graft failure 0.9415 vs. 0.8072, p = 0.005; rejection 0.7465 vs. 0.6413, p = 0.063). Moreover, MDs achieve slightly better results without CDSS. Finally, the table shows that using our cut-off (F2 for AI and > 0.5 for MDs) AI has got a higher sensitivity in comparison to MDs alone.

**Table 3:** AUC-ROC results on the reader study test set of AI, MD, and MD+AI for the endpoints death-censored graft failure and rejection. On the right side the table shows sensitivity (SEN) and specificity (SPE) with a cut-off of > 0.5 for MDs and F2 for AI.

| *Endpoint* | *Subject* | *ROC* | *SEN* | *SPE* |
| --- | --- | --- | --- | --- |
| *Rejection* | *AI* | **0.7465** | **0.5600** | 0.6947 |
|  | *MD* | 0.6413 | 0.4800 | 0.8316 |
|  | *MD+AI* | 0.6314 | 0.3200 | **0.8737** |
| *Graft Loss* | *AI* | **0.9415** | **0.6667** | **0.9247** |
|  | *MD* | 0.8072 | 0.5926 | 0.8387 |
|  | *MD+AI* | 0.7778 | 0.5926 | 0.8817 |

**Figure 2:**
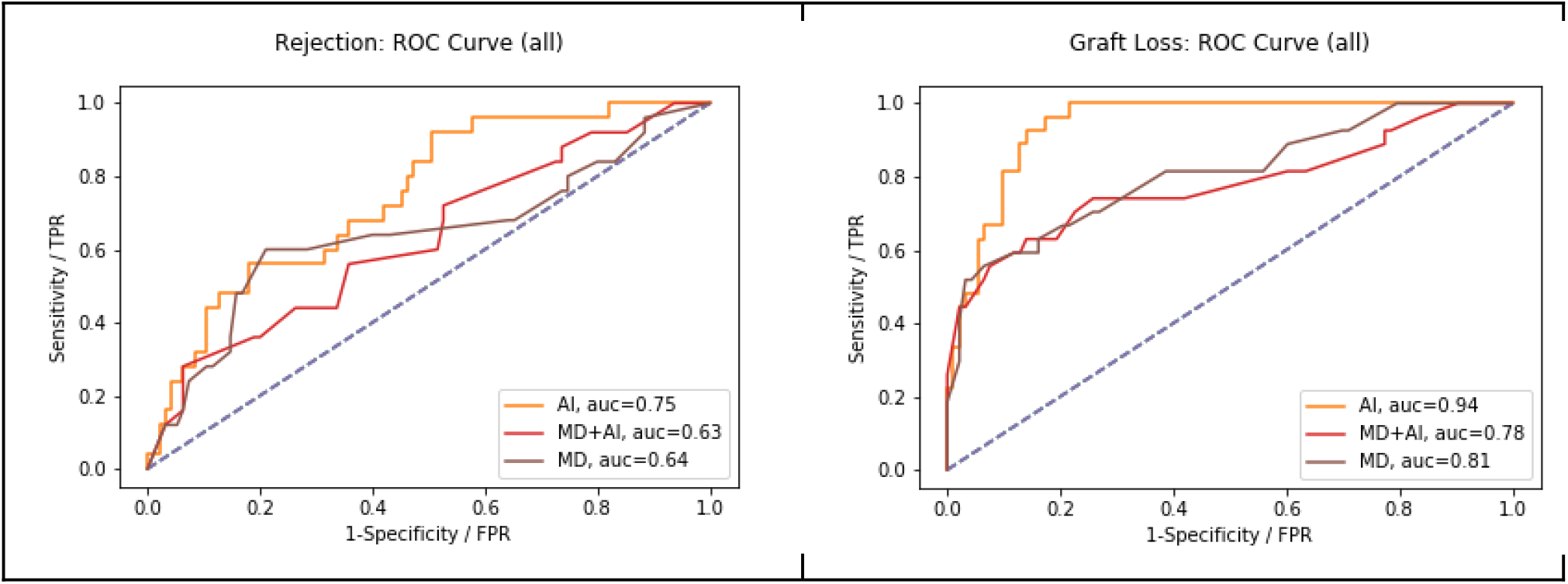
Performance of MD in comparison to AI and MD+AI, in terms of AUC-ROC on all three endpoints

Comparison of the results of junior MDs and senior MDs are presented in table 4. Without AI, senior MDs tend to score higher than junior MDs (graft failure 0.8108 vs. 0.8067; rejection 0.7134 vs. 0.5714). Moreover, junior MDs achieve higher results with CDSS for rejection (0.5714 vs. 0.7553, p = 0.075). Also, junior MDs with AI score higher than senior MDs with AI. And finally, senior MDs achieve a better score without AI.

**Table 4:** AUC-ROC performance of junior and senior physician groups in both parts of the study

| *Endpoint* |  | *Subject Groups* |  |
| --- | --- | --- | --- |
|  |  | *Junior MDs* | *Senior MDs* |
| *Rejection* | *MD* | *0.5714* | ***0.7134*** |
|  | *MD+AI* | ***0.7553*** | *0.4870* |
| *Graft Loss* | *MD* | *0.8067* | ***0.8108*** |
|  | *MD+AI* | ***0.8276*** | *0.7320* |

### Influence of the Clinical Decision Support System

To explore the influence of the CDSS on physicians, we calculate the difference of each prediction to the prediction made by the AI. MDs make a prediction in terms of probability (0-100%) and AI in terms of regression (0-100). Table 5 shows the mean difference scores, for all MDs as well as for junior and senior MDs separately. An additional box plot of the mean distance is presented in Figure 3. The table shows that in all cases the mean distance decreases if the physicians have access to the CDSS, and so does the standard deviation. Without the CDSS, physicians have on average a mean distance of 6 (overestimation of risk in comparison to AI). With the CDSS, the mean distance decreases in all cases, notably strong for junior MDs. Moreover, in the case of junior MD+AI, the mean distance to AI is below zero, which means that predictions are on average slightly below the score of the AI. In case of *all MDs* and *junior MDs*, the differences between MD and MD+AI are significant (p < 0.001). In the case of senior MDs we do not observe a significant difference. In addition to this, Figure 3 indicates that the variation of the predictions decreases with CDSS. In all three cases (MD+AI, junior MD+AI and senior MD+AI), more predictions are located closer to the prediction of the AI (located closer to the mean).

**Table 5:** Mean distance with standard deviation of estimation made by MD and MD+AI, to the prediction of the AI system. Distance score is calculated by subtracting the AI prediction from the human prediction. The lower line shows the 95% confidence interval (CI).

|  | *All MDs* | *Junior MDs* | *Senior MDs* |
| --- | --- | --- | --- |
| *MD* | 6.26 (31.35) | 7.99 (31.72) | 4.53 (31.02) |
| *MD+AI* | -0.06 (23.83) | -1.84 (24.05) | 1.73 (23.57) |
| *95% CI* | [1.75, 10.87] | [3.54, 16.12] | [-3.84, 9.44] |

**Figure 3:**
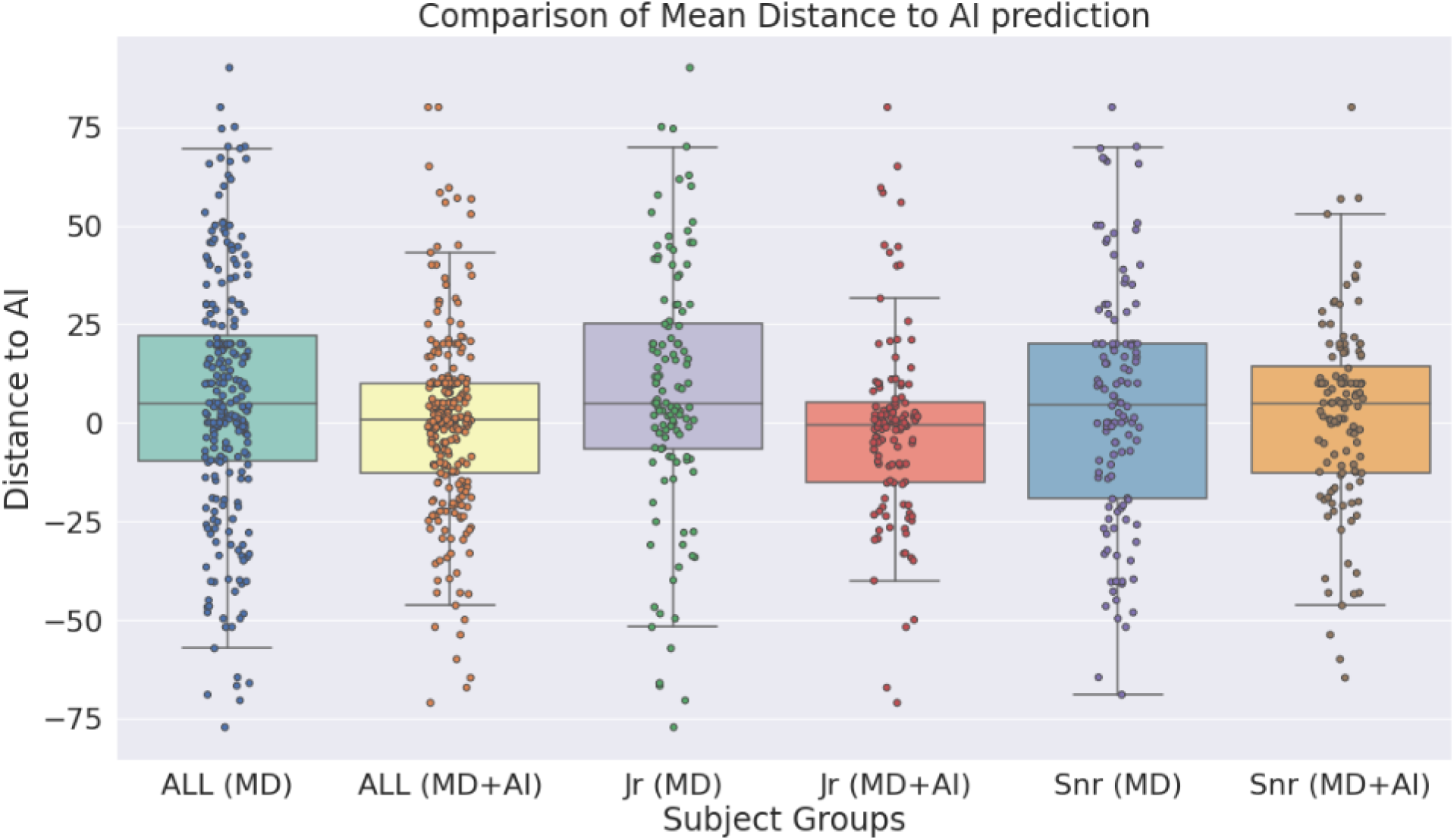
Box plot shows the distance of the estimations of medical doctors as well as medical doctors with clinical decision support system (CDSS), to the estimations of the CDSS alone.

### Accuracy of the predictions

The probability score of the physicians (cutoff = 50%) and the regression risk score (cutoff = F2-score; yellow and red zone in dashboard) were transformed into binary predictions: endpoint occurs, and endpoint does not occur (1/0).

Figure 4 presents the true/false positive predictions of the AI, as well as the physicians with and without CDSS. The figures show that in all cases some true positives (TPs) are not identified either by MD, AI, or MD+AI. Moreover, the figures present a large overlap, particularly for graft failure. Considering only MD and AI, the figure also shows that both make important contributions towards detecting many TPs. Even though AI scores higher in terms of ROC, it is not capable of detecting all critical cases, similar to MDs. Conversely, both together can detect more patients at risk beforehand, in comparison to MD or AI alone. For example, 7 Rejections were accurately predicted by both MD and AI, and additionally MD detected 5 rejections not detected by AI, and AI 7 rejections not detected by MD alone.

**Figure 4:**
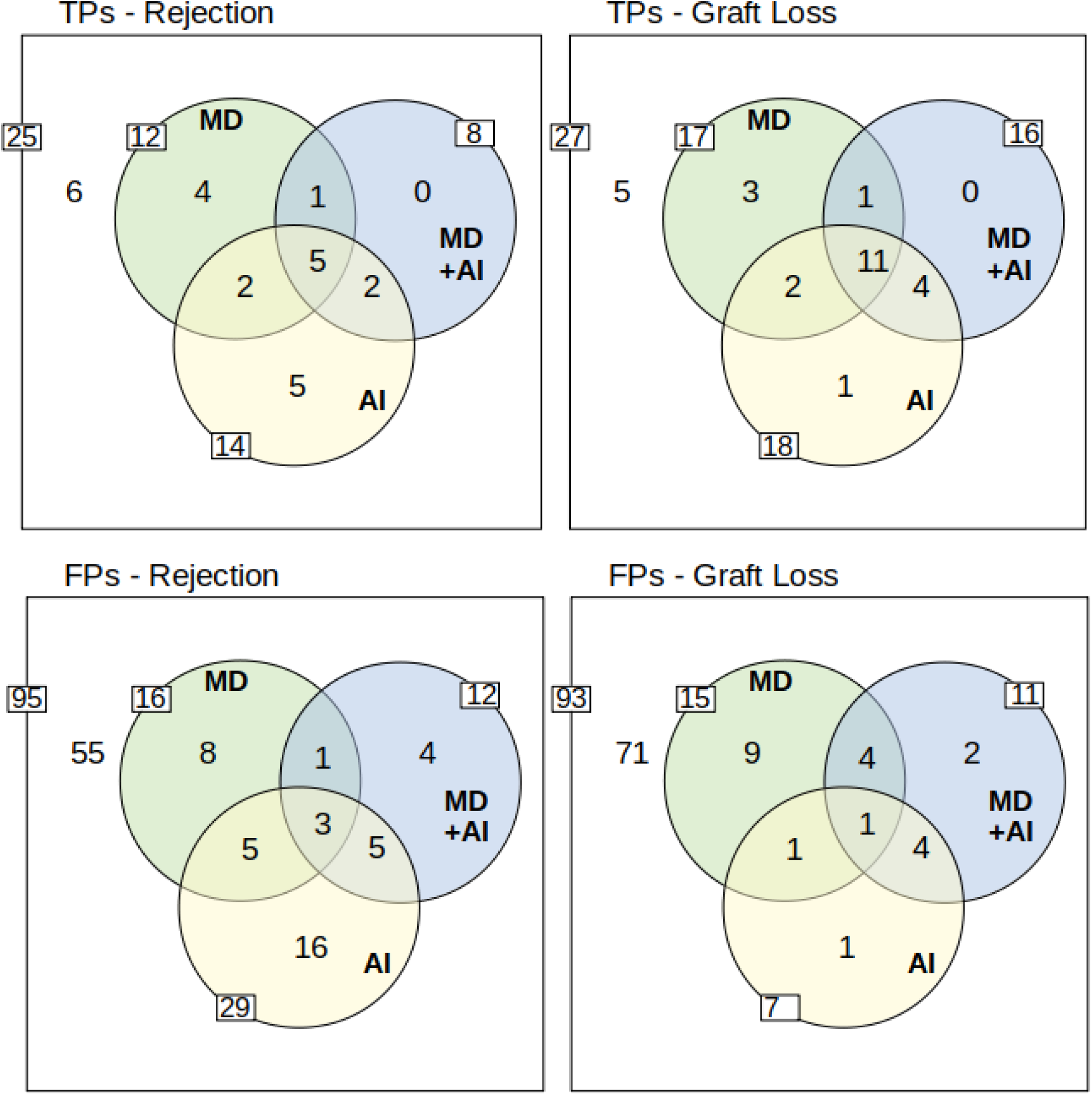
Overview of True Positives (TP) and False Positives (FP). The outer white rim describes the number of positives/negatives for each endpoint. The inner side of each circle indicates the number of true/false positives of AI, MD, and MD+AI. Moreover, the overlapping circles show the overlaps of true/false positive predictions between the different participants. The yellow circle of the AI system represents data points which were flagged with a yellow or red traffic light in the dashboard. To understand the example, take for instance TPs-Rejection: Overall 25 rejections (25 positives of 120 data points) occurred, from which 6 have not been detected by anyone in the study. AI predicted 14 (5+2+5+2) correctly, while MD predicted 12 (4+1+5+2), and MD+AI predicted 8 (1+5+2) correctly. 5 TPs are predicted by all (see intersection of all three circles). Moreover, two TPs are predicted only by MD and AI, and two other TPs only by MD+AI and AI. Only one TP is identified by MD and MD+AI. Finally, 4 TPs are predicted correctly only by MD, and 5 other TPs only by AI. MD+AI does not predict any additional TP which is already found by MD or AI. The lower row presents the same scenario for falsely predicted data points. A more detailed overview including also the results for the red and yellow warning of the traffic light system is included in the supplementary material.

The results indicate that MD+AI is oriented towards the AI system. Therefore, we can see a larger overlap between those two, and a smaller overlap to MD. AI was able to detect TPs that have not been identified by MD, and a reasonable amount of these is included in MD+AI. Yet, there are some TPs that have only been detected by AI alone. MD+AI did not lead to new TPs that had not been identified beforehand by either AI or MD.

As for false positives (FPs), AI has, except for graft failure, always the highest number of FPs. In case of rejection, AI makes 29 FP predictions, while the MDs makes only 16 and MD+AI only 12 FPs, respectively.

Table 6 shows positive predictive values (PPV) and sensitivity. The table shows that physicians alone can detect 48% of rejections and 63% of graft failures, while the PPV (precision) is 41% and 53%. The combination of both, MD and AI (MD U AI, not MD+AI), leads to an increased sensitivity, with a similar PPV level, in comparison to MD and MD+AI. For graft failure, 80% of the patients at risk can be detected, while only every second prediction is a FP - which is similar to the PPV of MD alone.

**Table 6:** Positive predictive value (PPV) and sensitivity (SEN) of MD and AI. The lower part of the table shows the union between MD and AI. Aligning the risk score to either zero or one, union means that we set the risk of a patient to one, if one of both identifies a potential risk.

| *Endpoint* | *Rejection* |  | *Graft Loss* |  |
| --- | --- | --- | --- | --- |
|  | *PPV* | *SEN* | *PPV* | *SEN* |
| *MD* | **0.41** | 0.48 | 0.53 | 0.63 |
| *AI* | 0.33 | 0.56 | **0.72** | 0.67 |
| *MD U AI* | 0.33 | **0.76** | 0.52 | **0.81** |

### Interpretation of false positives and false negatives

From a medical perspective, we observe that in case of FPs certain risk factors were present, and additionally that some endpoints occurred shortly after the target period of 90 days. Regarding FNs the retrospective second analysis revealed that no obvious known risk factors were present making it difficult to detect the endpoints for medical professionals.

### Endpoints predicted only by AI

Six endpoints are accurately predicted by AI but neither by MDs alone, nor by MD+AI. These data points are particularly interesting, as preventive measures, such as a closer monitoring could have avoided their occurrence if MDs would trust the CDSS enough to adjust the measures. Although difficult to objectively evaluate, from a medical perspective most of these cases are not obvious at first sight suggesting the potential for AI to detect additional kidney transplant recipients at risk.

## Discussion

Given the complexity of care after kidney transplantation and the diverse amount of potentially harming events, we evaluated a newly developed clinical decision support system (CDSS) for detection of patients at risk for rejection and graft failure in a reader study, in order to analyze assumptions made by physicians with and without the CDSS. First, our results shows that AI achieves better scores compared to physicians on our prediction tasks. Senior MDs perform better than junior MDs, but do not improve with the CDSS. Junior MDs with CDSS instead improve their capability to detect patients at risk. The fact that senior MDs alone perform better than junior MDs alone, might not be surprising. However, the reason why senior physicians have problems with the CDSS contrary to junior physicians remains unclear. One possible explanation is that junior MDs might be more open to new technologies. Also, as senior MDs have much more experience, they might have a stronger confidence in their opinion. This assumption might be supported by the fact that junior MDs converge stronger towards the suggestion of the CDSS. More studies are needed, to fully explore the reasons for these observations.

Another important result is that physicians and AI are able to detect different patients at risk. This highlights the potential benefit of a CDSS and provides strong arguments for its integration into clinical practice. At the same time, more research is needed on the appropriate mode of integration, given that AI resulted in many FPs and its recommendation did not improve senior MDs capabilities. Ultimately, multiple forms of explanations in a CDSS need to be explored and possibly adapted to the experience, the self-confidence and the affinity for technology of the different users.

Even though AI tends to outperform MDs, we need to keep in mind that ROC is just a score, which does not easily translate into a clinical benefit. When regarding sensitivity, specificity, and positive predictive value on the given task, we observe that physicians achieve higher sensitivity at the price of low specificity. One could conclude that MDs are more “cautious” than the AI used in this experiment. However, how can we benefit from such a system? The union of AI and MD could detect more critical patients - possibly by notifying the physician at the end of the treatment in case of a risk. Conversely, too many false alarms might also decrease the trust in such a system and might require too many resources^15^. To integrate such a risk prediction model into clinical care, more extensive studies are required. More reliable results might increase the trust in the new technology and lead to a better efficiency.

Our results are also relevant to a number of ethical issues surrounding AI-driven clinical decision support systems^16^. Besides debates on what it takes to trust medical AI and institutions deploying them^17^, and who bears moral and/or legal responsibilities for outcomes^18^, there are also opportunities: AI could play a part in empowering health systems, health institutions, and clinicians by making capacities widely accessible^19^. Our result that less experienced MDs improve their accuracy with the CDSS at hand provides a concrete example for such visions.

Similar to well-established diagnostic tests, or medical guidelines that are part of clinical routine, it might be possible that AI becomes an important part of evidence-based medicine in future. Thus, systems with high predictive power can even be perceived as having epistemic authority in their own right^20^: instead of AI being a mere decision aid, burdens of proof in case of deviation from system recommendations might shift to MDs, and individuals and institutions might systematically defer to them. Our results indicate that reality will likely be more fragmented for at least two reasons. First, since our system and MDs have different strengths, system-, user-, and context-specific understanding of these strengths will be essential towards potentially reaping the benefits of a “convergence of human and artificial intelligence”^21^ when seeking to optimize health outcomes. Second, the normative significance of various performance metrics is not definite and self-explanatory but requires continuous context-sensitive reflection and weighing involving different stakeholders. Besides, investigating the retrospective question, which factors caused false predictions, the prospective issue of what consequences a false prediction has for the patient will play a role in these discourses. For example, when predicting graft failure or rejection, the harms from FPs differ from those of FNs. Each way of privileging and weighing the metrics discussed reflects slightly different risk-benefit-tradeoffs, and high predictive power along one dimension, while promising and desirable, does not necessarily render the tool a gold standard for the task at hand.

While we use a large fine-grained German database collecting all routine clinical data for more than 20 years this approach has also some limitations. Ideally the model should be externally validated and may be adopted to a different data structure. It is important to highlight that the model should not be seen as an universal “out of the box” model. Instead, it provides a good “baseline” model, which can be improved or may provide the basis for a lean model that is easy to implement in different contexts. However, these issues are beyond the scope of this article, where we focused on the human-machine interaction that can influence future design and implementation of ML-models in kidney transplant care.

## Supporting information

TRIPOD checklist

## Data Availability

Data are available on reasonable request from the corresponding author.

## Disclosures

The authors of this document have no conflicts of interest to disclose.

## Author Contributions

RR: planning, technical development, paper writing; MM: conceptual design, conduction of the study, paper writing, technical implementation; BO: conduction of the study, paper writing; DS: technical implementation & technical support; KB conceptual design, paper writing; All authors contributed to discussion and reviewed the paper.

## Acknowledgement

This project was partially funded by the Federal Ministry of Education and Research (BMBF, Germany) in the project “vALID: Al-Driven Decision-Making in the Clinic. Ethical, Legal and Societal Challenges” (No 01GP1903B), and by the European Union’s Horizon 2020 research and innovation program under grant agreement No 780495 (BigMedilytics). Any dissemination of results here presented reflects only the author’s view. The European Commission is not responsible for any use that may be made of the information it contains”. Dr. Marcel Naik is a participant in the BIH Charite Digital Clinician Scientist Program funded by the Charité – Universitätsmedizin Berlin, the Berlin Institute of Health and the German Research Foundation (DFG).

## Supplementary Material Table of Content

A. Endpoints

B. Data

C. Data Selection, Enrichment and Cohort Generation

D. Pre-Processing and Feature Extraction

E. Cohort Generation - Exclusion Criteria

F. Dashboard

G. Internal Validation

H. Analysis of Prediction

I. Metrics

J. Power analysis

K. Instructions for participants

## Notes

### Competing Interest Statement

The authors have declared no competing interest.

### Author Declarations

Ethics committee of Charité - University Hospital Berlin gave ethical approval for this work.

