## Supplementary Data for "Evaluation of a clinical decision support system for detection of patients at risk after kidney transplantation"

**Supplementary Material**

As the main article is restricted in size, many details about our work, particularly about the data and the model, are presented in this supplementary material.

A. **Endpoints**

**Death-Censored Graft Failure** This endpoint is defined as the initiation of renal replacement therapy (dialysis or re-transplantation) due to transplant failure, which is well documented. It is important to highlight that graft loss due to death with a functioning graft is not included.

**Rejection** Since our center’s policy calls for exclusion of potentially reversible factors by biopsies in all patients with elevated serum creatinine levels, which are not sufficiently explained by other reasons, such as urinary tract obstruction or infection, we defined rejection according to the Banff 2017 classification^1^, which has been re-read and re-assessed by two independent nephropathologists as previously described^2^. Endpoint definition criteria are met in case of Banff diagnostic category 2 “Antibody-mediated changes” and/or Banff diagnostic category 4 “TCMR”.

B. **Data**

Baseline for this work is TBase, a database designed for kidney transplant recipients (KTR) and candidates, implemented over 20 years ago at Charité - Universitätsmedizin Berlin, including >6000 patients, >7500 kidney transplantations, >200000 diagnoses, >54000 medical reports and >24 million laboratory values reference^3^. As patients are supposed to receive a follow-up at the transplant center 3-4 times a year, TBase includes fine-grained information about the patients over many years including demographics, laboratory data, medication, medical notes, diagnoses, radiology, and pathology reports. Additionally, TBase includes information on hospitalisation in the transplant center as well as discharge diagnoses (ICD10 codes) and procedures (OPS codes).

Overall, the quantity and quality of the data has improved over time since its introduction in 1999. Moreover, some fields of the database allow a free text input, which makes it sometimes difficult to automatically process data. Also, within each follow-up different data can be collected or different checks can be done - depending on specific requirements for the patients.

C. **Data Selection, Enrichment and Cohort Generation**

The complete risk prediction scenario is built up around data points, which describe the moment when new data about a patient is inserted into TBase. In our case, a data point relates to follow-ups (triggers e.g. vital signs), lab values, and hospitalizations. For each data point of each patient, we select all patient information, which is available at that point in time. Moreover, we examine how many days after that an endpoint occurs (if at all). If the endpoint occurs within our target prediction window of 90 days, it will be labelled as **true**, otherwise as **false**. In addition to that, we enrich the data by information, which will also be used as additional features in our model, such as mean scores or gradients of successive values.

Next, data is filtered to generate a meaningful, reliable, valid and realistic dataset. First, pediatric KTR (<18 years at transplantation) had been excluded. Then, data points in which an endpoint is active are removed. In addition, data points less than one week after the diagnosed endpoint rejection are excluded, as the endpoint still might be active. Furthermore, the first two weeks after a transplantation are dismissed, as lab values might have a strong variation within that period. Finally, only data points with a follow-up datapoint within the next 15 to 180 days are used. This filter has been implemented in order to exclude gaps in patient follow-up ensuring reliability of endpoint evaluation. An overview about this filtering is provided in Table S1.

In the rest of this work, we refer to this resulting dataset as “cohort”. As presented in Table S3, this cohort consists of 102918 different data points and the majority of the data points have a negative outcome (no endpoint). Table S2 presents some basic patient characteristics of the cohort. For instance, the cohort includes 1516 different patients (925 male, 590 female), with a mean of 67.89 data points per patient. Overall, 211 patients suffered at least once a rejection, 199 patients at least once a graft loss, and 79 of those patients suffered both, at least one rejection and at least one graft loss. Considering the last data point of each patient, the mean distance to the last transplantation is about 8 years (2948 days). Moreover the table shows the mean value (of the mean of each patient) of four crucial lab values.

D. **Pre-Processing and Features**

To convert the cohort into a training, development, and test dataset for ML, data has to be cleaned up and features extracted. As various fields of the database can be noisy, significant effort has been put into improving the quality of the data. This included for instance typos in diagnosis text fields, different measure units for the same lab values, as well as the parsing and removal of “false” entries, such as, e.g., fields of lab values, which include characters in numeric fields. In addition to that, plural forms and synonyms in the diagnosis were normalized, and single tokens used as binary word features. Note, no imputation has been applied, as preliminary experiments did not show promising improvements.

For each endpoint a different model has been trained. Overall we have got a large range of different features consisting of structured information such as medications, ICD10 codes, or lab values, as well as bag-of-word features extracted from a text field defining the underlying renal disease. Particularly for values such as lab values and vital parameters, we consider the last two occurring values. Moreover, additional features have been generated from the given features as follows:

- **Day Distance**: For each follow-up, hospitalization, and lab value the distance (in days) of given entry and the reference datapoint is calculated.
- **Mean Values**: Within a period of one year (before datapoint), we calculate for various lab values the mean of that patient.
- **Above/Below**: For the last two lab results (inducting a range of different values) we calculate the distance to the mean value.
- **Past Endpoints**: We store how often our target endpoints occurred within the last 180, 360 and 1800 days.
- **Gradient**: We calculate the gradient between the last two vital measurements, distance divided by number of days.
- **Past Examinations**: We store how often an analysis of lab results was requested in the last 60, 120 and 240 days.

In this way, a pool of about 1200 different features were generated. However, each model (which relies on a combination of 300 pruned trees of max depth of 3), utilises about 300 different features, which are summarised in Table S1, and presented in detail in Table S6.

E. **Cohort Generation - Exclusion Criteria**

Figure S1 depicts how the cohort is created, and particular data points excluded.

F. **Dashboard**

Figure 1 (main document) presents an example of the dashboard, presenting the time on the x-axis and the risk score on the y-axis. All data points of the last year, including corresponding risk scores, are visualized in the graph. The graph itself is divided into different zones, a green zone indicating a low risk that the target endpoint will occur, a yellow zone which indicates a higher risk, and a red zone which indicates the highest risk. Each zone is defined by a threshold, which was generated on the development set by identifying the optimal F-Scores - F2 score for the border to the yellow zone, and F0.5 to indicate the red zone.

On the right side, the number of TPs (true positives) in the given zone are presented, as seen in the development set. This information helps the physician to understand limitations of the system in a better way, as, e.g., not all data points in the red zone are necessarily TPs.

In the lower part of the dashboard, two different explanations are provided. The lower left side presents features with a strong influence on the current score (local features). The values are identified by applying an ablation, re-testing the model and removing different features each time. The visualized features indicate those, which lead to the highest drop in performance after removal. The drop of performance is expressed as “relevance”. The lower right side presents features which are in general relevant for the overall model (global features). Features are identified using gini importance, which takes into account how many nodes of the trees a feature occurs in - the more frequent the more important.

G. **Internal Validation**

Table S4 presents an alternative to Table 2 (internal validation) in the main document, presenting sensitivity and specificity using the F2 and F0.5 thresholds of the traffic light system, as well as their confidence interval (CI) on the 50 repeated experiments.

H. **Analysis of Prediction**

In comparison to Figure 4 (main document), Figure S2 presents a much more detailed overview about the true/false positive predictions of the AI system, as well as the physicians with and without AI support, in comparison to “Analysis of Predictions” in the main document. In the figures, the outer white rim describes the number of positives/negatives for each endpoint. The inner side of each circle indicates the number of true/false positives of AI, MD and MD+AI. Moreover the overlapping circles show the overlaps of true/false positive predictions between the different participants. The yellow/red circle of the AI system represents data points which were flagged with a yellow/red traffic light in the dashboard.

I. **Metrics**

In the following we define the different metrics which have been used, whereas *true positives* are defined as TP, *true negatives* as TN, *false positives* as FP and *false negative* as FN:

**Sensitivity** (also known as *true positive rate*, and *recall*) is defined as:

Sensitivity = TP / (TP+FN)

**Specificity** (also known as *true negative rate*) is defined as:

Specificity = TN / (TN+FP)

**Positive predictive value** (*PPV*, also known as *precision*) is defined as follows:

PPV = TP / (TP+FP)

**F-Score** is a joint score calculated from precision (PPV) and recall (sensitivity). The most commonly used score is the F1-Score, which describes the harmonic mean between precision and recall. In this work however, we use the F2-Score, which weights recall higher than precision, and the F0.5-Score, which weights precision higher than recall. F2 and F0.5 are used to define the traffic light system (yellow and red area), applied to the development set to find the threshold. The F-Score is defined as follows, whereas ß has to be filled with the corresponding value (e.g. 1, 0.5 or 2):

F$\beta$ = (1+$\beta$^2^) * (precision * recall) / (( $\beta$^2^ * precision) + recall )

J. **Power analysis**

Power calculation was performed before conducting the reader study, using the method of Konietschke and Brunner.^4^ We used the following assumptions: AUC-ROC of MD is 0.6 and AUC-ROC of MD+AI is 0.8, since AI achieved AUC-ROC values between 0.8 and 0.95 during internal validation. When for 60 out of 120 datapoints used in the reader study, at least one endpoint occurs (20 graft loss, 20 biopsy-proven rejection, 20 infection based on CRP definition), the power is 0.84. Alternatively, for AUC-ROC of MD 0.7 and AUC-ROC of MD+AI 0.85, the power is 0.71, and for AUC-ROC of MD 0.7 and AUC-ROC of MD+AI 0.9, the power is 0.96.

K. **Instructions for participants**

Original instructions were given in German. We provide an English translation without modifications in terms of content or form.

**K1) Procedure**

You have a maximum of 30 minutes to study a patient, whose data from the database TBase are displayed pseudonymized until a predefined “censoring” point in time (Illustration 1).

After 30 minutes, or by clicking on “Finish”, you are asked to enter your estimation of the probability in percent at which a biopsy-proven rejection episode, or graft failure (see Table SK1 for definition) will occur within the next 90 days. Additionally, you are asked to justify the respective decision-making. Afterwards, the next patient is displayed. The risk probability entered should be on a scale from 0 to 100, where 100 means that the event occurs with absolute certainty. Values below 50 would mean that the end point would "rather not" occur. Conversely, values over 50 mean the end point is “more likely” to occur. The data points were not selected entirely random, indicating that the distribution of the endpoints does not have to correspond to reality.

In the first part (physician without AI), you will evaluate a total of 15 patients.

In the second part of the experiment (physician with AI), you will evaluate 15 different patients, but will be provided risk predictions of an AI tool.

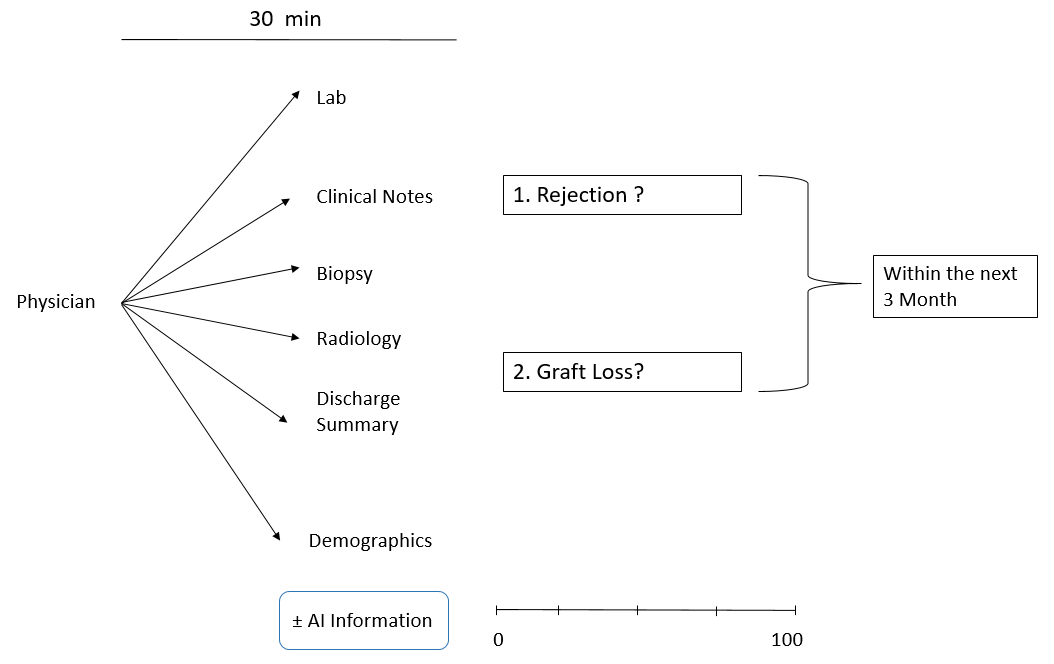

Figure SK1: Study summary for participants.

| **Endpoint** | **Definition** |
| --- | --- |
| Rejection | Rejection Defined via Banff 2017 classification (biopsy necessary)  ● Category 2: “Antibody-mediated changes”  AND OR  ● Category 4: “TCMR” |
| Graft failure (Death-censored graft failure) | ● Initiation of renal replacement therapy  ● Excluding death with a functioning graft |

Table SK1: Definition of the endpoints for the participants

**K2) AI model - Basic information information for participants**

| Model | Gradient Boosted Regression Trees: Put simply, this is a tree-based process that combines a variety of mini-trees. In our case a model with 300 trees is used. Trained on data, it can recognize patterns and correlations to make predictions on unseen data. |
| --- | --- |
| Data | The model is based on data from Charité Mitte and Virchow Klinikum between 2008-2020. Mainly, demographic data, laboratory, diagnoses, medications and information about transplantation are taken into account. The data are very “unbalanced” with respect to the endpoints. That means that there are many more data points, for which in the next 90 days the endpoint does not occur than for which it actually occurs. The exact ratio is shown below under “Bias”. |
| Filtering | The following data points were filtered out of the data before the development and experiment:  - data from patients under 18 years of age;  - data points for which one of the endpoints is currently active;  - all data points that appear within a period of up to one week after a rejection;  - all data points that occur up to two weeks after transplantation date  - all data points from patients, which do not have a follow-up data point in the next 15-180 days |
| Evaluation | The current models were evaluated by a 50-fold cross-validation, whereby individual patients with their respective complete data, in training, validation and test set have been divided. |
|  | Graft Failure Rejection |
| Performance | ROC-AUC ROC-AUC  0.945 0.832 |
| Explanation | Put simply, the ROC-AUC score provides information on how well “positive” and “negative” events can be differentiated. Are positive events rated rather high and negative events tend to be rated rather low? A random sort would achieve a ROC-AUC score of 0.5, the maximum with a perfect test is 1.0. The ROC-AUC score is very popular, but in the case of severely unbalanced data, it can still show a high score if there are many false positives. |

**K3) Visualization of the ML-based risk prediction:**

- For every patient, and for every endpoint, there is a single graph.

- A score between 0-100 is provided.

- In addition, the value is shown on a traffic light system (green, yellow, red)

- For example, green has a lower probability that the endpoint occurs, than yellow or red.

- The corresponding probability of occurrence was determined on so-called validation data (not the test data used in this experiment)

- Risk probabilities for a specific score differ from endpoint to endpoint

- The “risk progression” of the previous data points (if available) of the patient are displayed in the same graph. The graph does not show the values of data points that were filtered out before.

- Relevant features (variables) for the system’s decision are displayed in the graph of Figure SK2.

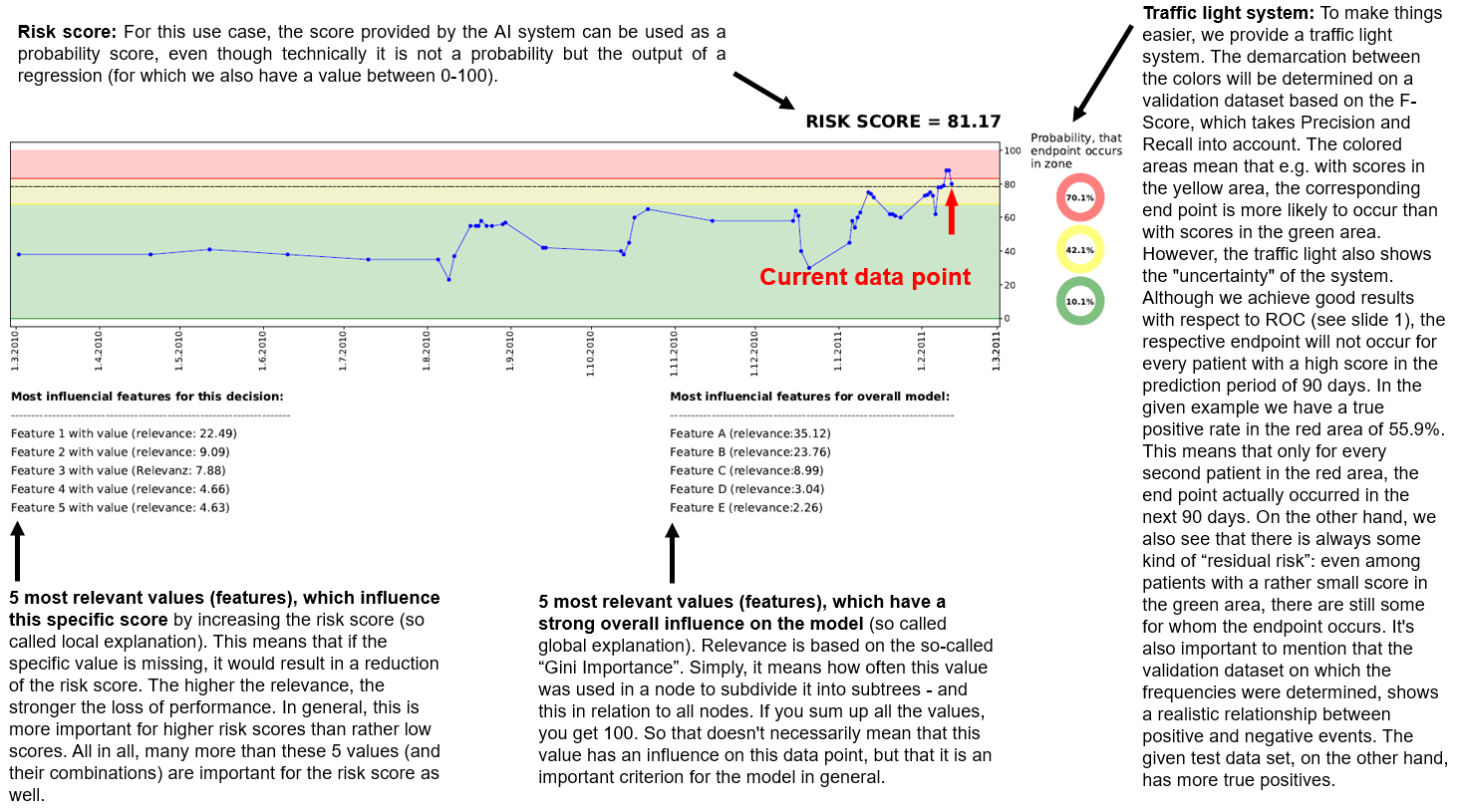

Figure SL2: Detailed description of the visualization of the ML-based risk prediction for participants.

***Supplementary Figures***

***Figure S1:***

*Cohort generation, including exclusion criteria and number of data points.*

***Figure S2:***

*Overview of True Positives and False Positives. To understand the example, take for instance* ***TPs-Rejection****: Overall 25 rejections (25 positives of 120 data points) occurred, from which 6 have not been detected by anyone in the study. AI predicted 14 correctly, while MD predicted 12, and MD+AI predicted 8 correctly. 5 TPs are predicted by all participants, but only one, if the red traffic light is used to classify predictions, instead of the yellow one. Moreover, two TPs are predicted only by MD and AI, and two other TPs only by MD+AI and AI. Only one TP is identified by MD and MD+AI. Finally 4 TPs are predicted correctly only by MD, and 5 other TPs only by AI. MD+AI does not predict any TP which cannot be found by MD or AI. The lower row presents the same scenario for falsely predicted negative data points.*

***Figure S1:***

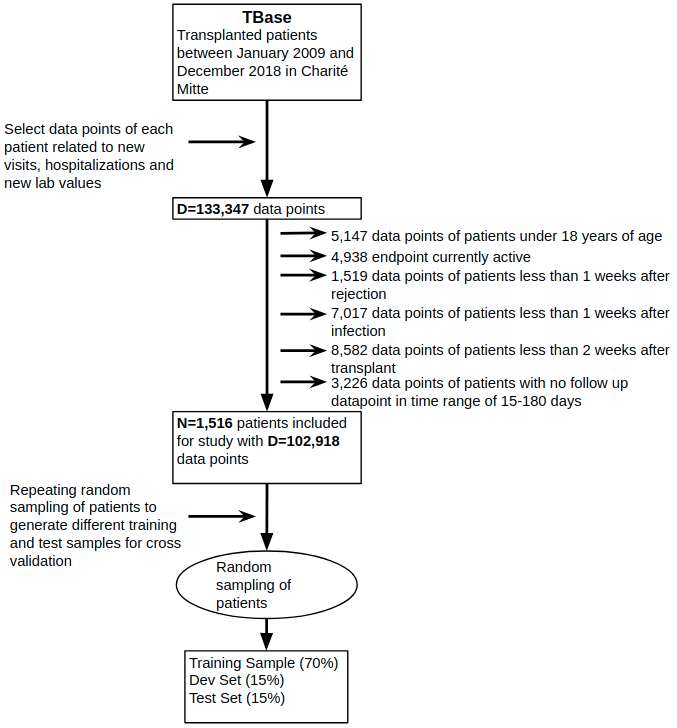

***Figure S2:***

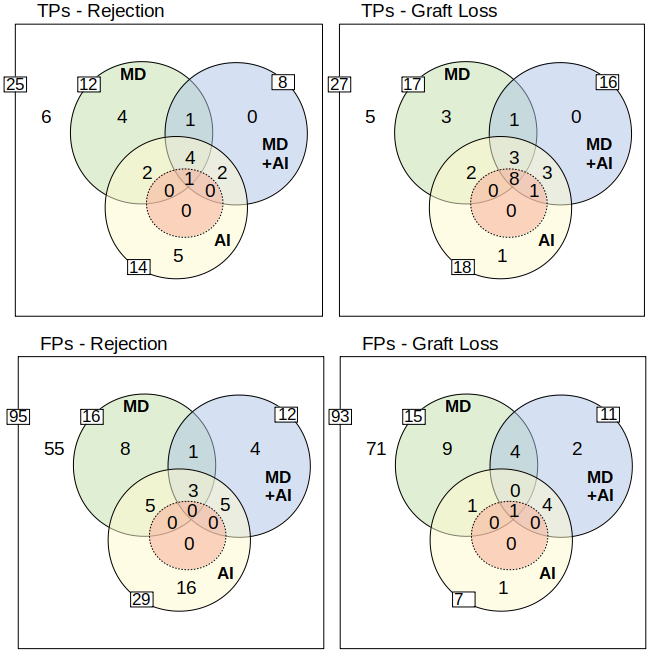

***Supplementary Tables***

***Table S1:***

*Summarised overview about the features used in each model, as well as the number of different features considering both models together.*

| ***Feature Category*** | ***Rejection*** | ***Graft Loss*** | ***Both*** |
| --- | --- | --- | --- |
| ICD10 Codes | 120 | 147 | 205 |
| Lab Values | 100 | 85 | 106 |
| Medications | 17 | 32 | 40 |
| Bag-of-Words (BoW) | 21 | 18 | 34 |
| Analytics Features | 19 | 9 | 19 |
| Transplant | 8 | 8 | 10 |
| Vital Parameters | 7 | 7 | 7 |
| Patient | 5 | 4 | 5 |
| Overall | 297 | 310 | 426 |

***Table S2:*** *Analysis of relevant patient information of given cohort*

| ***Demographics*** |  |
| --- | --- |
| *Number of Patients:* | *1516* |
| *Age (mean/std)* | *54.77 / 16.54* |
| *Male/Female/Other* | *925 / 590 / 1* |
| *Days since transplantation (mean/std)* | *2947.95 / 2228.46* |
| ***Endpoints per Patient:*** |  |
| *Rejection* | *211* |
| *Graft Loss* | *199* |
| *Overlap* | *79* |
| ***Lab Values (mean/std):*** |  |
| *creatinine (mg/dl)* | *2.53 (1.37)* |
| *Leuko (/nl)* | *8.04 /nl (2.25)* |
| *eGfr (ml/min)* | *41.69 (17.75)* |
| *Hb (g/dl)* | *11.72 (1.40)* |

***Table S3:***

*Frequency of positive and negative events across all endpoints.*

|  | ***Rejection*** | ***Graft Loss*** |
| --- | --- | --- |
| #pos / #neg | 2827 / 100091 | 2677 / 100241 |
| ratio | 1:35 | 1:37 |

***Table S4:***

*Results from Table 2 (main document) for the estimation of 90 days, mapped to Sensitivity and Specificity, using F2 (traffic light: yellow) and F0.5 (traffic light: red) as threshold.*

| ***Endpoint*** | ***Threshold*** | ***Sensitivity*** | ***Specificity*** |
| --- | --- | --- | --- |
| ***Rejection*** | *F0.5 (red)* | *0.227, 95% CI [0.190, 0.265]* | *0.965, 95% CI [0.954, 0.977]* |
|  | *F2 (yellow)* | *0.519, 95% CI [0.490, 0.549]* | *0.882, 95% CI [0.869, 0.895]* |
| ***Graft Loss*** | *F0.5 (red)* | *0.273, 95% CI [0.249, 0.297]* | *0.993, 95% CI [0.992, 0.994]* |
|  | *F2 (yellow)* | *0.724, 95% CI [0.703, 0.746]* | *0.952, 95% CI [0.949, 0.956]* |

***Table S5:*** *Participant baseline characteristics. Mean age, clinical experience and experience in kidney transplant care in years for junior physicians (MD) and senior MD, respectively.*

| ***Participant***  ***Group*** | ***Number female/male*** | ***Age***  ***(mean)*** | ***Clinical experience (mean)*** | ***Experience in kidney transplant care (mean)*** |
| --- | --- | --- | --- | --- |
| ***Junior MD*** | 3 / 1 | 32 | 3.5 | 2.5 |
| ***Senior MD*** | 2 / 2 | 41 | 13.5 | 8.75 |

***Table S6:***

*Detailed overview about the different features of each model*

| ***Feature Category*** | ***Feature Name*** | ***Rejection*** | ***Graft Loss*** |
| --- | --- | --- | --- |
| ICD10 | *z94.0*  *z92.2*  *z92.1*  *z90.5*  *z86.7*  *z51.4*  *z29.0*  *z03.8*  *y57.9*  *t86.19*  *t86.12*  *t86.11*  *t86.10*  *t85.71*  *t83.8*  *t82.8*  *t81.8*  *t81.4*  *t81.3*  *r80*  *r55*  *r50.9*  *r11*  *q61.9*  *q61.3*  *q44.6*  *n40*  *n30.0*  *n26*  *n25.8*  *n19*  *n18.84*  *n18.83*  *n18.5*  *n18.3*  *n18.2*  *n18.0*  *n17.91*  *n17.9*  *n17.8*  *n13.3*  *n13.1*  *n05.9*  *n04.9*  *n04.2*  *n04.1*  *n03.3*  *m81.9*  *m54.4*  *m51.2*  *m34.0*  *k76.0*  *k52.9*  *k35.9vlz*  *k29.6*  *k29.5*  *j99.1*  *j96.00*  *j81*  *j18.9*  *j15.8*  *i95.1*  *i82.88*  *i70.20*  *i69.3*  *i51.9*  *i50.13*  *i50.12*  *i49.0*  *i48.10*  *i47.1*  *i42.0*  *i36.1*  *i35.1*  *i35.0*  *i34.0*  *i31.3*  *i27.28*  *i25.22*  *i25.19*  *i25.11*  *i15.11*  *i15.10*  *i15.1*  *i12.9*  *i10.01*  *i10.00*  *i10*  *i07.1*  *h35.3*  *h35.0*  *h26.8*  *g62.9*  *g62.88*  *f43.2*  *e89.0*  *e87.2*  *e86*  *e83.58*  *e78.0*  *e75.2*  *e53.8*  *e13.90*  *e11.90*  *e11.9*  *e11.21*  *e11.20*  *e05.8*  *e04.9*  *e03.9*  *d90*  *d63.8*  *d50.8*  *c61*  *a46*  *a41.9*  *a09.0*  *a09*  *a04.7*  *- (no code)*  *z99.2*  *z95.5*  *z88.8*  *z49.1*  *z22.3*  *z09.80*  *z09.0*  *z03.5*  *t86.9*  *t86.82*  *t86.1*  *t83.5*  *t82.7*  *t82.5*  *t82.3*  *r50.80*  *r10.4*  *r10.3*  *r06.0*  *n39.0*  *n32.8*  *n25.0*  *n18.82*  *n18.4*  *n17.99*  *n17.92*  *n13.5*  *n12*  *m31.3*  *k92.2*  *k83.0*  *k74.6*  *k65.0*  *k57.32*  *k57.30*  *k56.6*  *k43.9*  *k40.90*  *j98.1*  *j96.0*  *j44.11*  *j18.8*  *j18.1*  *j06.9*  *i73.0*  *i70.22*  *i63.4*  *i48.9*  *i48.19*  *i42.2*  *i33.0*  *i27.0*  *i25.13*  *i25.12*  *i21.4*  *i20.9*  *i20.8*  *i12.00*  *i11.00*  *i10.91*  *i10.90*  *g63.2*  *g47.31*  *e87.5*  *e83.5*  *e79.0*  *e78.5*  *e78.2*  *e21.3*  *e21.1*  *e11.72*  *e10.9*  *e10.72*  *e10.20*  *d64.8*  *d62*  *d37.4*  *c83.3*  *c64*  *c44.3*  *c44.2*  *b99*  *b18.2*  *b02.9*  *a41.0* | x  x  x  x  x  x  x  x  x  x  x  x  x  x  x  x  x  x  x  x  x  x  x  x  x  x  x  x  x  x  x  x  x  x  x  x  x  x  x  x  x  x  x  x  x  x  x  x  x  x  x  x  x  x  x  x  x  x  x  x  x  x  x  x  x  x  x  x  x  x  x  x  x  x  x  x  x  x  x  x  x  x  x  x  x  x  x  x  x  x  x  x  x  x  x  x  x  x  x  x  x  x  x  x  x  x  x  x  x  x  x  x  x  x  x  x  x  x  x  x | x  x  x  x  x  x  x  x  x  x  x  x  x  x  x  x  x  x  x  x  x  x  x  x  x  x  x  x  x  x  x  x  x  x  x  x  x  x  x  x  x  x  x  x  x  x  x  x  x  x  x  x  x  x  x  x  x  x  x  x  x  x  x  x  x  x  x  x  x  x  x  x  x  x  x  x  x  x  x  x  x  x  x  x  x  x  x  x  x  x  x  x  x  x  x  x  x  x  x  x  x  x  x  x  x  x  x  x  x  x  x  x  x  x  x  x  x  x  x  x  x  x  x  x  x  x  x  x  x  x  x  x  x  x  x  x  x  x  x  x  x  x  x  x  x  x  x |
| Lab Values | *Standard bicarbonate*  *Urine sodium > 1 mmol/l*  *Serum sodium*  *TSH*  *Transferrin saturation*  *Transferrin*  *Tested prothrombin time*  *INR*  *Thrombocyte count*  *Triglyceride level*  *Tacrolimus level*  *Total urine volume (24 hour urine collection)*  *Standard bicarbonate (2)*  *Specific gravity dipstick urine*  *Reticulocyte count*  *Reticulocyte hemoglobin*  *Red cell distribution width*  *Parathormon*  *Proteinuria (quantification 1)*  *Dipstick proteinuria*  *Proteinuria (quantification 2)*  *Serum protein*  *Proteinuria (quantification 3)*  *Proteinuria (quantification 4)*  *pH dipstick urine*  *Serum phosphorus*  *Blood pH*  *Procalcitonin*  *Blood oxygen saturation*  *Neutrophil count*  *Serum sodium*  *Urine sodium concentration*  *Mean platelet volume*  *Monocyte count*  *Serum magnesium concentration*  *Urine magnesium conctration*  *Mean corpuscular volume*  *Mean corpuscular hemoglobin*  *Mean corpus hemoglobin concentration*  *Lymphocyte count*  *Relative lymphocyte count*  *Serum lipase*  *Leukocyte dipstick urine*  *Leukocyte count*  *Leukocyte urine microscopy*  *LDL cholesterol*  *Lactate dehydrogenase*  *Urine creatinine concentration*  *Serum creatinine concentration*  *Urine creatinine concentration (2)*  *Urine creatinine concentration (3)*  *Creatinine clearance*  *Body size (necessary for clearance estimation)*  *Body weight (necessary for clearance)*  *Serum potassium*  *Urine potassium*  *Urine potassium (2)*  *Immature granulocyte count*  *Relative immature granulocyte count*  *Serum urea concentration*  *Urine urea concentration*  *Serum uric acid concentration*  *HLA luminex class 2 screening test*  *HLA luminex class 1 specifying test*  *HLA luminex class 1 screening test*  *Hematokrit*  *HDL cholesterol*  *Serum hemoglobin concentration*  *HbA1c*  *Glucose dipstick urine*  *Serum glucose concentration*  *Serum glucose concentration*  *Gamma GT*  *eGFR*  *eGFR (2)*  *Ferritin*  *Everolimus level*  *Erythrocyte count*  *Erythrocyte urine microscopy*  *Eosinophil count*  *Relative eosinophil count*  *Iron concentration*  *Ciclosporin level*  *C-reactive protein*  *Creatine kinase*  *Cholesterol*  *Serum calcium*  *Blood dipstick urine*  *Total bilirubin*  *Direct bilirubin*  *Basophil count*  *AST*  *Activated partial thromboplastin time*  *Alkaline phosphatase*  *ALT*  *Albuminuria*  *Albuminuria (1)*  *Serum albumin*  *Albuminuria (2)*  *Albuminuria (3)*  *Serum osmolality*  *Urea (other material)*  *Haptoglobin*  *Serum chlorine*  *Bilirubin dipstick urine*  *Conjugated bilirubin* | x  x  x  x  x  x  x  x  x  x  x  x  x  x  x  x  x  x  x  x  x  x  x  x  x  x  x  x  x  x  x  x  x  x  x  x  x  x  x  x  x  x  x  x  x  x  x  x  x  x  x  x  x  x  x  x  x  x  x  x  x  x  x  x  x  x  x  x  x  x  x  x  x  x  x  x  x  x  x  x  x  x  x  x  x  x  x  x  x  x  x  x  x  x  x  x  x  x  x  x | x  x  x  x  x  x  x  x  x  x  x  x  x  x  x  x  x  x  x  x  x  x  x  x  x  x  x  x  x  x  x  x  x  x  x  x  x  x  x  x  x  x  x  x  x  x  x  x  x  x  x  x  x  x  x  x  x  x  x  x  x  x  x  x  x  x  x  x  x  x  x  x  x  x  x  x  x  x  x  x  x  x  x  x  x |
| Medications | *ranitidin* | x |  |
|  | *natriumbicarbonat* | x |  |
|  | *mycophenolatmofetil* | x |  |
|  | *metoprolol* | x | x |
|  | *methylprednisolon* | x | x |
|  | *metamizol* | x |  |
|  | *levothyroxin-na* | x |  |
|  | *lercanidipin* | x | x |
|  | *epoetin beta* | x | x |
|  | *darbepoetin alfa* | x | x |
|  | *ciprofloxacin* | x |  |
|  | *ciclosporin* | x | x |
|  | *carvedilol* | x |  |
|  | *calcitriol* | x | x |
|  | *benzbromaron* | x | x |
|  | *amlodipin* | x |  |
|  | *acetylsalicylsäure* | x | x |
|  | *valsartan* |  | x |
|  | *valganciclovir* |  | x |
|  | *torasemid* |  | x |
|  | *tacrolimus* |  | x |
|  | *pantoprazol* |  | x |
|  | *omeprazol* |  | x |
|  | *nph-insulin* |  | x |
|  | *natriumhydrogencarbonat* |  | x |
|  | *mycophenolatnatrium* |  | x |
|  | *kaliumdihydrogenphosphat/natriummonohydrogenphosphat* |  | x |
|  | *kaliumcitrat* |  | x |
|  | *kalium* |  | x |
|  | *folsäure* |  | x |
|  | *fluvastatin* |  | x |
|  | *enalapril* |  | x |
|  | *doxazosin* |  | x |
|  | *digitoxin* |  | x |
|  | *dalteparin* |  | x |
|  | *colecalciferol* |  | x |
|  | *calciumcarbonat* |  | x |
|  | *benazepril* |  | x |
|  | *azathioprin* |  | x |
|  | *allopurinol* |  | x |
| BoW | *wegener*  *the*  *streptococcal*  *schoenlein_henoch*  *proliferativ*  *post*  *nephrosklerose*  *nephropathie*  *nephronophthise*  *nephritis*  *mesangiale*  *membranous*  *interstitial*  *iga*  *hydronephrose*  *glomerulonephritis*  *diabetes*  *cirrhosis*  *chronisch*  *amyloidose*  *- (no entry)*  *unspecified*  *tumor*  *sponge*  *segmentale*  *pyelonephritis*  *polyzystische*  *of*  *membranoese*  *membrano-proliferative*  *kidney*  *hypertensive*  *focal*  *cystinose* | x  x  x  x  x  x  x  x  x  x  x  x  x  x  x  x  x  x  x  x  x | x  x  x  x  x  x  x  x  x  x  x  x  x  x  x  x  x  x |
| Analytics Features | *Analytics_xDiff*  *Has currently AKI Risk*  *Has currently AKI Failure*  *Had reject in last 360 days*  *Had reject in last 180 days*  *Had reject in last 1800 days*  *Had infection in last 180 days*  *Had infection in last 1800 days*  *Days since last vital parameter*  *Number of lab values (last 60 days)*  *Number of lab values (last 240 days)*  *Number of lab values (last 120 days)*  *Number of lab examinations (last 60 days)*  *Number of lab examinations (last 240 days)*  *Number of lab examinations (last 120 days)*  *Days since last lab value*  *Days since last hospitalization*  *Number of stations in hospital*  *Number of days in hospital* | x  x  x  x  x  x  x  x  x  x  x  x  x  x  x  x  x  x  x | x  x  x  x  x  x  x  x  x |
| Vital Parameters | *urine volume*  *temperature*  *heart frequency*  *weight*  *diuresis time*  *blood pressure systolic*  *blood pressure diastolic* | x  x  x  x  x  x  x | x  x  x  x  x  x  x |
| Transplant | *Donation type (living vs. deceased)*  *Primary function*  *Number of transplanation*  *Mismatch grade (split)*  *Mismatch grade (broad)*  *Time since last transplantion (months)*  *Cold ischemia time*  *EBV IgG*  *HCV antibodies*  *Dialysis type* | x  x  x  x  x  x  x  x | x  x  x  x  x  x  x  x |
| Patient | *rh factor*  *body size*  *gender*  *blood type*  *age* | x  x  x  x  x | x  x  x  x |

problems in small sample sizes. *Computational Statistics & Data Analysis, 53*(3), 730-741.
